# Artificial intelligence and health equity in primary care: A qualitative study with key stakeholders

**DOI:** 10.1101/2023.10.25.23297533

**Authors:** Alexander d’Elia, Mark Gabbay, Lucy Frith, Sarah Rodgers, Ciara Kierans

**Author notes:** Corresponding author: Dr Alexander d’Elia, 3^rd^ Floor, Whelan Building, University of Liverpool, Brownlow Hill, Liverpool, L69 3GB, United Kingdom.

## Abstract

Artificial Intelligence (AI)-augmented interventions are currently being rolled out across primary care, but how it affects health equity remains insufficiently understood. This qualitative study addresses this gap through an ethnographical inquiry based on 32 interviews and focus groups with stakeholders including commissioners, decision makers, AI developers, researchers, GPs and patient groups involved in the implementation of AI in English primary care. We took a sociotechnical perspective in order to assess how the stakeholders can improve health equity through the implementation process of AI within the wider system. We found that regulation and policy alone cannot guarantee equitable implementation of AI but can provide a framework to enable other stakeholders to take measures to promote equity: fostering a shared understanding of the causal mechanisms of AI and health equity, how to measure health equity, and how to share data necessary for equity promotion. Further, all stakeholders need to be on board for equitable implementation, and currently innovation leaves clinicians and patients behind. Capacity building is needed to achieve this, in particular at local commissioning and clinician level. Careful implementation and pragmatically focused research are needed to make AI in primary care capable of advancing health equity.

## Introduction

Artificial intelligence (AI) can be described as a computer system that performs tasks traditionally requiring human intelligence. Everyday examples include predicting preferences in social media feeds, recognizing faces in photos, or recently, compiling complex information in natural language such as ChatGPT ^1^. A rapidly expanding field, AI-augmented interventions are considered integral to the healthcare of the future and current applications include interpreting X-rays and ECGs. Widespread application of AI-based innovations ^2^ in healthcare is already happening, and primary care is no exception.

At the same time, health inequity (hereafter *inequity*) is increasingly discussed, not least in the context of the ongoing COVID-19 pandemic ^3^. Through potentially freeing up resources and enabling more personalized care, AI has been described as a potential enabler for more equitable healthcare ^2^. However, as AI interacts with socioeconomic, gender and ethnic health inequities on many different levels, it could both increase or decrease inequities, depending on application and implementation ^4^ ^5^. Primary care holds a unique role in tackling inequity. It can be both a source and a magnifier of inequities, as well as a platform for mitigation ^6^. For the purpose of this study, primary care is defined as primary care services provided to individual patients, specifically in the English National Healthcare System (NHS), and not including wider public health policy ^7^.

In the context of inequity and implementation, AI differs from other digital interventions in a number of ways. The most important of these are: in contrast to other complex health-informatics interventions; AI-augmented interventions are likely to be more opaque in showing how outputs are achieved ^8^; AI has the potential (largely untapped at the moment) to be self-improving, raising issues around evaluation ^9^; AI opens up new domains of healthcare provision to digitalization and privatization, which may have unforeseen effects on the primary-care system and on health equity ^10^ ^11^. Finally, the pace of innovation brought on by AI is unprecedented in healthcare, further elevating long-standing concerns about technology implementation in healthcare (e.g. poor adaptation and adverse system effects ^12^) ^13^.

This study is underpinned by a comprehensive literature review describing the pathways for how AI can affect inequity in primary care ^14^, producing a framework for how AI may affect inequity *in and through* primary care (Appendix 1). Our review found a multitude of potential mechanisms, broadly divided into ‘intrinsic’ and ‘extrinsic’ effects. Intrinsic effects include inequity impact from skewed, or biased, data collection and algorithms that may contribute to the different quality of care different population groups might receive (e.g., Obermeyer et al ^15^). Extrinsic effects on the other hand take place through the implementation of a given AI-augmented intervention and the interaction between the AI-intervention and the system it is deployed in. A typical example is the ‘digital divide’; the tendency for socioeconomically disadvantaged groups and the elderly to have less access to technology and thus benefitting less from AI interventions requiring such technology compared to their more advantaged peers. Our review also discussed potential extrinsic inequity issues stemming from a lack of patient trust, increased focus on self-management (the success of which correlates to socioeconomy), adding workload to already strained clinical workforce causing out-crowding effects, and a general risk that interventions target uncomplicated, easy patients over patients with complex socially multimorbidity. If these inequity-exaggerating effects are to be avoided or mitigated, and conversely if the potentially equity-positive effects are to be gained, careful implementation of AI products is necessary, which in turn requires a thorough understanding of the system in which they are being deployed. The importance of taking this social context into account when implementing health technology has been extensively studied. For example a 2017 meta-analysis found that sociotechnical factors were involved in more than two thirds of health technology interventions that failed to achieve their objectives ^12^ ^16^.

There are numerous reports on how to best implement AI in healthcare and other sectors, typically by governmental and non-governmental organisations ^2^ ^17–22^. Being typically general and (intentionally) speculative they provide a starting point for further research, but few provide empirically grounded findings. Among the more ambitious of such projects is a recent report by The Ada Lovelace Institute, ‘The Knotted Pipeline: Data-driven systems and inequities in health and social care’ ^23^. The report builds on interviews with organizational stakeholders in the health-data context (including but not exclusively AI) and concludes that there is a distinct need for a common definition of equity and for improved collaboration between different organisations in order to improve health equity outcomes thorough health data. The report also emphasizes the perils of overestimating the ability of data-driven technologies in improving outcomes when they are being implemented in an existing, highly complex system; the risk of viewing AI as a panacea and failing to take the wider organisational and social environment into account. In contrast to the Ada Lovelace piece, we focus on AI and primary care specifically, apply a different and specific research approach for data collection and analysis, and involve a diverse set of stakeholders, including patients. To summarize, our study aimed to assess how the sociotechnical network in which UK primary-care AI-implementation is taking place (i.e., the organizations, regulations and stakeholders involved) is conducive to making AI a force for health equity, and what makes for a favourable environment for reducing health inequity.

## Materials and Methods

This is a qualitative, explorative, social-constructivist study drawing primarily from 27 individual interviews and five focus groups with stakeholders involved in the implementation of AI in primary care.

### Participants

The selection of participants was both purposive and theory driven. The purposive sampling meant a broad representation of different stakeholders, whilst the theory-driven aspect allowed for continuous recruitment as the study went on with interviewees being recruited to explore emerging themes. In practice this meant deciding early on which broad groups were seen as integral stakeholders and thus important to include (the purposive aspect), but also to draw on the findings from the interviews and recruit sequentially to explore the themes that were developing (the theory-driven aspect). For example, early interviews showed that algorithmic impact assessment (AIA) were a key component, and thus people with specific experience of this were recruited (e.g., #17 from the Ada Lovelace Institute). Participants consisted of:

- *Developer* stakeholders. Product developers working on primary-care AI products. Recruited by contacting a range of UK AI companies involved in primary care via public email addresses.
- *General Practitioners* (GPs) Recruited by disseminating the invitation through South Birmingham GP Training Scheme mailing list, and through snowballing from interviewees.
- *Commissioning* stakeholders focusing on Integrated Care System (ICS) and Academic Health Service Networks (AHSN) members involved in digital product procurement. Recruited through emailing all ICSs and AHSNs across England, of which a minority were available for interviews.
- *Regulator* stakeholders representing the organizations central in implementation process, including the Medicines and Healthcare products Regulatory Agency (MHRA), the National Institute of Care Excellence (NICE), the Department of Health and Social Care (DHSC), the Care Quality Commission (CQC) and the Information Commissioners Office (ICO), as well as the advisory non-governmental Ada Lovelace Institute. Recruited through emailing respective organization.
- *Researcher* stakeholders (i.e., researchers investigating topics of AI and its impact on healthcare), including both clinical academics as well as non-medical researchers. Recruited through targeted emails and subsequent snowballing.
- *People living with diabetes mellitus* (insulin- and non-insulin dependent) (PwDs). Recruited via Diabetes UK’s West Midlands office, who in turn asked local organizations to help with the recruitment of participants for focus group sessions during their local meetings around the West Midlands, England. PwDs were recruited as a case study for patients with chronic illnesses in general, and the emphasis was on the participatory component of their own healthcare and their relationship with primary care and AI.

To capture divergent ideas and reach adequate fidelity for every interviewed group, it was decided that every stakeholder group would have at least five interviewees. However, only four commissioning stakeholders were interviewed, due to a last-minute cancellation.

### Inclusion and exclusion criteria

Participants from the groups representing their professions/organisations were not selected based on demographic characteristics. No individual demographic selection of PwDs included was made, however PwD groups were purposively sampled to an extent as the research question necessitated diverse representation (a minimum of 1/3 of either gender, and 1/5 of minority ethnicity for all PwDs). Exclusion criteria for all participants included:

- Lacking capacity to consent to participation.
- Not proficient in English (due to pandemic-related limited resources of clinically confidential interpretation services it was not feasible to offer this).

### Methodology

This study applied an iterative constant-comparative methodology, as described by Silverman ^24^, allowing for an effective method for conducting explorative research from a series of sequential interviews ^25^. Analysis was conducted using three levels of coding in an iterative manner, inspired by Grounded Theory ^26^; herein referred to as codes, themes and theory (technically a middle-range theory).

Initially, the implementation systems and processes were mapped in general terms using publicly available documents and complemented by interview data as well as a conceptual model of equitable implementation (appendix 2 and 3, respectively), to provide a backdrop for the rest of the study. The collection and analysis of the interview data thereafter consisted of six steps, repeated until saturation (Figure 1). All steps were conducted by the first author with advisory support from the co-authors, apart from step 3:

1. Data collection: interviews and focus groups. All data collection sessions were conducted online using Zoom videocall software ^27^, with the exception of three of the PwD focus groups, which were conducted at local Diabetes UK meetings. Diabetes UK sessions were not recorded, but field notes were taken. Both interviews and focus group sessions were semi-structured, with different interview schedules depending on what themes were being explored. Transcription was conducted using Sonix software ^28^ with subsequent manual editing. Anonymization took place after transcription with each transcript given a code (Table 1) and recording deleted. Data was then imported into NVivo 12.0 qualitative analysis software for subsequent steps ^29^.
2. Familiarization; a read through of the data without coding. Comments were made based on free association.
3. Initial, free coding, of transcripts followed by comparison with all the previous codes to decide if a new code should be created or the excerpt should be coded under an existing code. The coding was predominantly conducted by the first author, but two transcripts were double coded by CK and MG to improve internal validity.
4. Codes were then collated into themes. As with the initial codes, all new themes were compared to the previous themes avoid duplication, all existing themes were reviewed in each round, and altered if needed to remain compatible with all initial codes. (See appendix 4 for examples of iterations of the theme list).
5. Building provisional middle-range theory from themes; the theory acted as a glue that related the themes to each other, keeping the inquiry focused on the research question. The theory was revised continuously in line with the development of the themes.
6. Recruit new participants to explore the themes and theory further; develop interview guides on new lines of inquiry. Again, this recruitment was partially guided by the theory, partially by the criteria of including all stakeholder groups.

**Figure 1:**
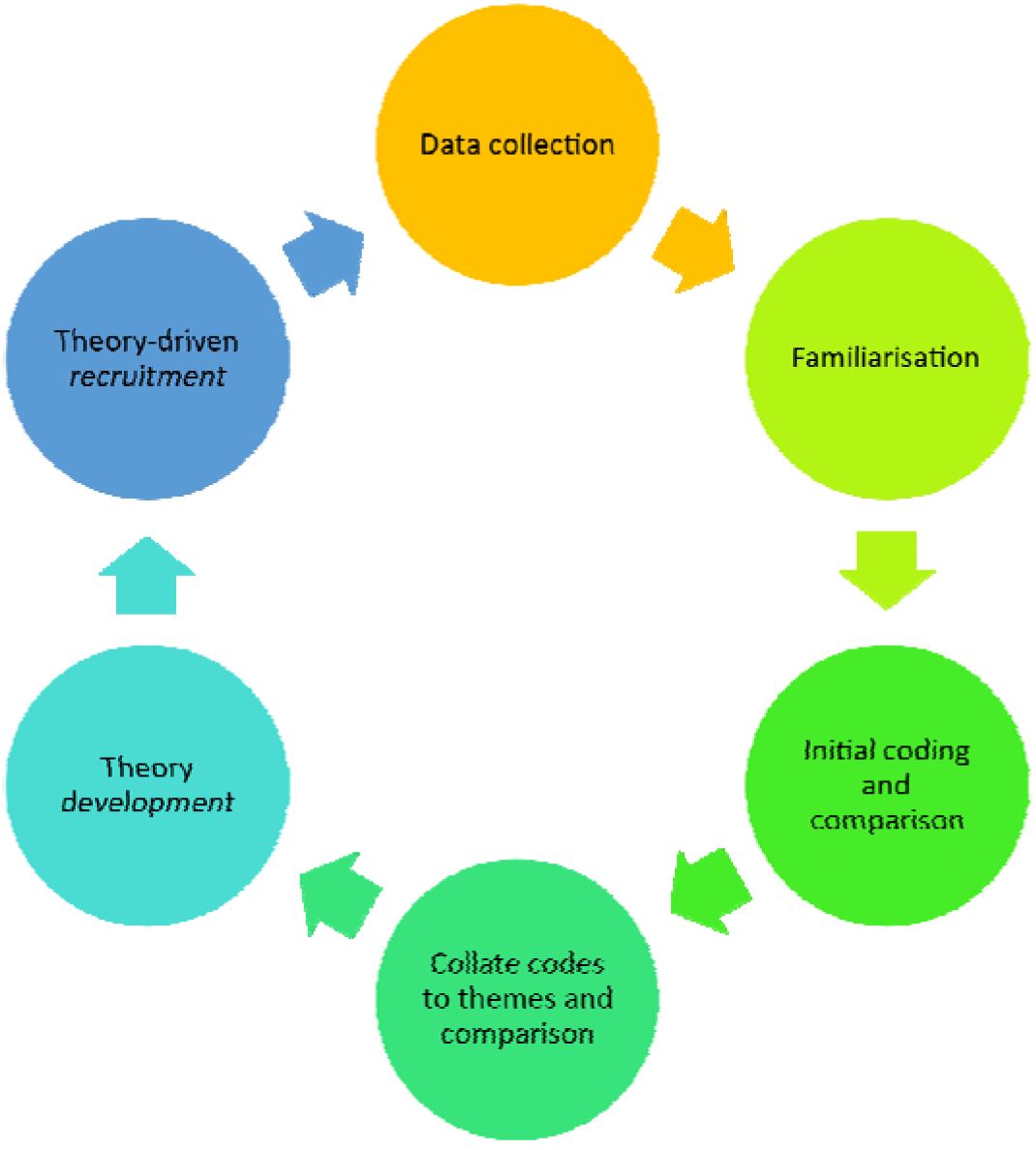
The iterative process for data analysis and recruitment. 1) Data collection, 2) Familiarization of transcripts and field notes, 3) Initial coding and comparison, 4) Collate codes into themes. 5) Build provisional advance theory from themes. 6) Recruit new participants to explore the themes and theory further. Reiterate as new data are collected until saturation.

**Table 1:** Overview of individual interviews and group interviews with people living with diabetes (PwDs). Note that age and ethnicity were only recorded for the latter. Numbers refer to anonymization code, broadly corresponding to the order of recruitment.

| Professional stakeholders | % female | Professional stakeholders | % female |
| --- | --- | --- | --- |
| Total subjects (excl. focus groups) n=27 | 36 % | Commercial developers n=4 | 25 % |
|  |  | 19 |  |
| General Practitioners n=7 | 57 % | 20* |  |
| 1 |  | 22 |  |
| 2 |  | 23 |  |
| 3 |  | Commercial developers: Focus group n=1 | (33 %) |
| 5 |  | 21 (n=6) |  |
| 7 |  | Local commissioners (ICS) and<br>Advanced Health Service Networks<br>(AHSN) n=4 | 50 % |
| 11 |  | 6 (ICS) |  |
| 16 |  | 8 (ICS) |  |
| Regulatory/advisory stakeholders n=6 | 33 % | 13 (AHSN) |  |
| 4 (Dept. Of Health and Social Care) |  | 15 (AHSN) |  |
| 10 (Medicines and Healthcare products<br>Regulatory Agency) |  | Researchers n=6 | 17 % |
| 12 (Information Commissioners Office) |  | 9 |  |
| 17 (The Ada Lovelace Institute) |  | 14 |  |
| 18 (Care Quality Commission) |  | 24 |  |
| 27 (National Institute for Health and<br>Care Excellence) |  | 25 |  |
|  |  | 26 |  |
|  |  | 28 |  |
| PwDs: Focus groups n=4 | % female | % >65 years | % minority ethnicity |
| PwD 1 (n=7) | 57 % | 100 % | 29 % |
| PwD 2 (n=6) | 33 % | 83 % | 50 % |
| PwD 3 (n=6) | 67 % | 100 % | 0 % |
| PwD 4 (n=10) | 60 % | 70 % | 10 % |
| Total PwDs: n=29 | 55 % | 86 % | 21 % |
| Average duration of interviews: 39 minutes (range 23-63). Focus groups: 43 minutes (33-51) |  |  |  |

These steps were repeated as new data were collected, until saturation.

Finally, the results were compiled as a narrative, building on the final iteration of the themes and the joining theory, and compared to preceding scholarship. A graphical logic model serves to visualize the theory (Figure 2), which has been used in studies with similar methodology ^30–32^.

**Figure 2:**
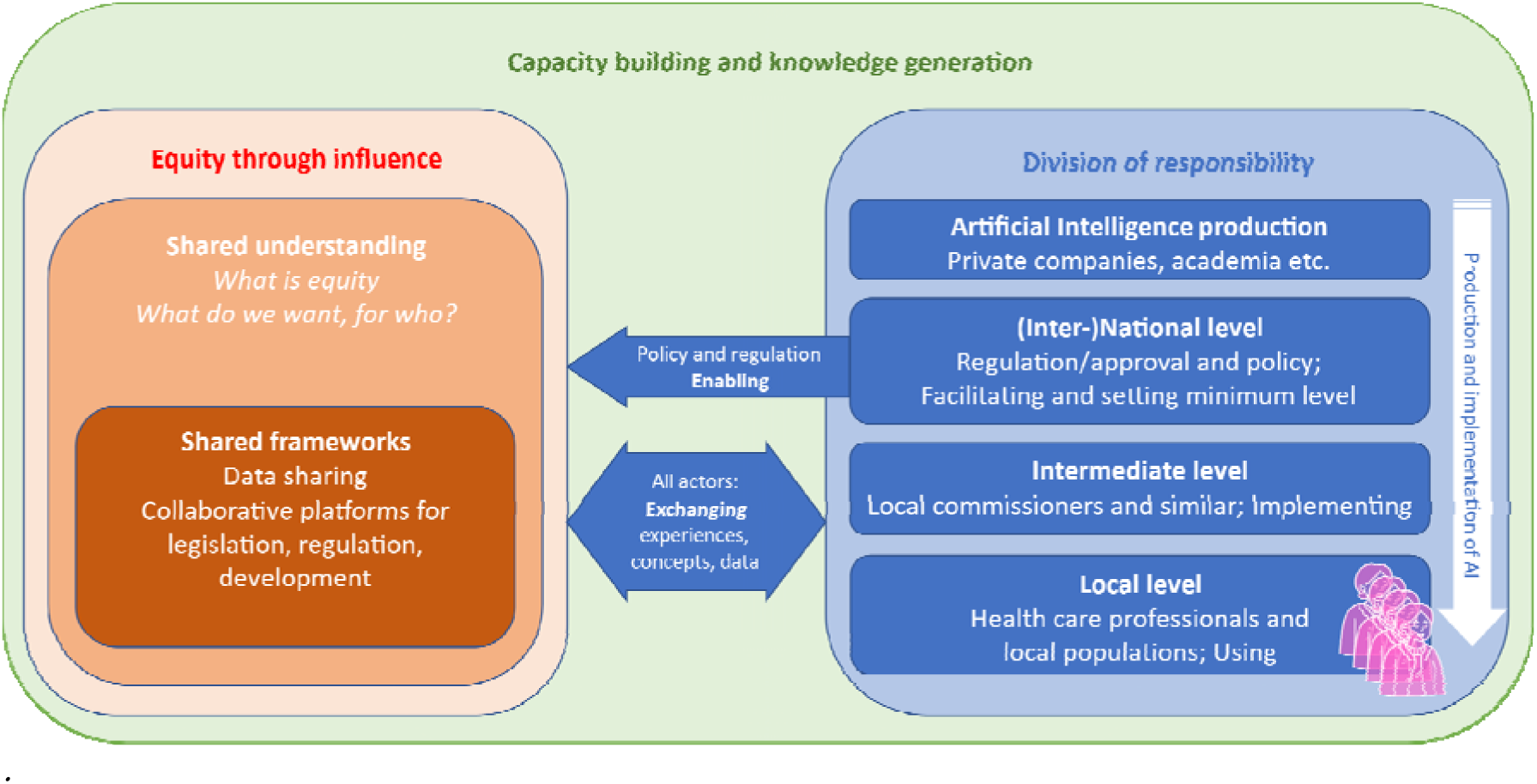
The theory: the concluding model for equitable implementation of AI in primary care.

### Public involvement

Patient and/or public involvement (PPI) in research and development and implementation of medical AI has been suggested as a way of improving trust and reducing the risk of adverse health-equity effects ^15^ ^33^, as has been previously discussed in non-AI contexts ^34^ ^35^. In the context of medical AI, previous research has shown a link between PPI to improve public understanding of AI and acceptability of a hypothetical AI product, predicting that this would lead to more successful implementation ^36^.

During the development and carrying out of this study, two remunerated public advisors were involved, helping to shape the direction of the study, the interpretation of the results (i.e., the Discussion section) and the dissemination. The advisors identify with traditionally marginalized populations (one of British Asian ethnicity and one registered disabled and member of the LGBT community) and helped widen the perspectives of the inquiry. On a larger scale, the two advisors have been involved in the planning of the PhD project this study forms part of, providing input on useful areas of inquiry, overarching methodology and dissemination.

### Reflexivity

The first author is a general practitioner in training in the English NHS. Subsequently, the study has been conducted from the perspective of a practical primary care practitioner. It is likely that the findings would have been slightly different if a commissioning stakeholder or private AI company had conducted a similar study, due to differences of perspective. Second, the main author is a White, 34-year-old, healthy, medically educated male; a privileged group in terms of inequity. This clearly affects understanding of inequity, even as the same has considerable clinical and academic experience of inequity. The two public advisors’ background helped broaden perspectives.

### Ethics

This study was approved by The University of Liverpool, research ethics committee, reference number 10229. The study was conducted in accordance with the recommendations for physicians involved in research on human subjects adopted by the 18th World Medical Assembly, Helsinki 1964, and later revisions.

## Results and Interpretation

### Demographics

Twenty-seven individual interviews and five focus group sessions (all PwD sessions plus one of the commercial developer sessions, #20) were conducted during ten months (April 2022 to January 2023). Of these, GPs were the largest group, with seven interviews conducted, followed by researchers (n=6), regulatory and advisory stakeholders (n=6), commercial developers (n=4 plus 1 focus group), local commissioners (n=4) and PwD focus groups (n=4). All these sessions were conducted remotely. Demographic information was not collected for the professional stakeholders (i.e., all but PwDs) beyond gender, which ranged from 17 % female (researchers) to 57 % female (GPs). For all professional stakeholders 36 % were female. The average duration of the interviews was 39 minutes (ranging from 23 to 63 minutes).

The four PwD groups ranged in size from six to ten participants with a total of 29. All but one of these sessions (PwD 4) were conducted in person at the respective Diabetes UK meeting location. For PwDs basic information on age and ethnicity was collected to meet the diversity criteria of the study. Of all PwD participants, 55 % were female (33-67 %), 86 % were over 65 years old (70-100 %) and 21 % were of non-White-British ethnicity ^37^ (0-50 %). The average focus group lasted for 43 minutes (33 to 51 minutes).

### Addressing the aim

Our aim is to to assess how the sociotechnical network in which UK primary-care AI-implementation is taking place (i.e., the organizations, regulations and stakeholders involved) is conducive to making AI a force for health equity.

Comparing the data and derived themes and joining theory (Figure 2) with the mentioned model of equitable implementation (appendix 3), a complicated picture of the current sociotechnical network emerges. Many of the actions indicated as required in this ‘ideal’ model indeed appear to take place; algorithmic impact assessments are developed to assess the impact of AI in various settings (although no primary care specific ones exist), initiatives to improve information flow between different stakeholders are taking place (e.g. the AI and Digital Regulations Service/ADRS and AHSNs). Arguably, looking beyond AI, this push for local involvement is one of the reasons for the recent reorganization of local NHS groups into larger regional Integrated Care Systems.

Our interview and focus group data showed that more needs to be done for the system to be conducive to equitable implementation. The five themes discussed in this section represent five interconnected approaches to ‘catalyze’ such an implementation chain, and the above graphical logic model (Figure 2) – the *theory* – illustrates how they relate to each other.

#### Theme 1: Equity through influence

Starting from the left side of the theory model (Figure 2), *equity through influence* encompasses the inherent power imbalance between the different stakeholders when shaping a new intervention, and AI interventions in particular. Extensively studied and described in other healthcare contexts ^38–40^, equitable implementation of technology needs to be addressed across the hierarchy of stakeholders, simultaneously. Applying an implementation science perspective ^41^ leads to the conclusion that in order for an intervention to function well (and in the context of the NHS this includes promoting equity), an intervention must be anchored in the behavior of all its stakeholders, and the communication culture between these stakeholders. As one interviewee involved in an advisory organization put it:

> I think there needs to be a much more sophisticated public dialogue around applications of AI and health, the public need to be joined in with it. The government needs to be participating in this conversation, and it shouldn’t be consultative [i.e., not just a one-time exercise]. We shouldn’t just throw out something and say, this is happening, deal with it. There needs to be sustained, meaningful public engagement in this space. I think that is the number one takeaway from my work.
>
> #17, Ada Lovelace Institute

Consequently, ‘bringing all stakeholders on-board’ and making sure that the intervention responds to all stakeholders’ needs throughout both development, implementation and evaluation is essential. Involving the public (patient and public involvement/PPI) is needed to understand local needs and local inequity:

> I think a lot of it will come down to the creators [of AI products] to have a greater understanding of the needs of the population that they’re aiming for. And that’s how you can make things and bespoke to the context. I think some of these companies must actually go into primary care to see what happens at the reception to kind of see what the queries are from patients and stuff.
>
> #1, GP

A PwD expressed frustration over how new interventions in primary care did not seem to be in the interests of the patients:

> All that is needed is some common sense. If they knew how we experienced [living with diabetes] they would be able to [provide care] better!
>
> PwD, group 1

However, both in the context studied in this project and in general, there is a lack of consensus on how to best conduct PPI for improved equity in outcomes ^42^, which needs to be addressed. Participatory primary care clinics, such as the Good Things Foundation’s NHS Digital Health Labs ^43^ offer an interesting approach. This initiative, which ran between 2017 and 2020 in the South of England, included ‘Digital Health Hubs’; open spaces where community members could interact with the care system (e.g., their primary care practice) with various digital tools. As part of this they were studied and interviewed, both in the moment and as structured focus groups, and the mainly focusing on accessibility of digital user interfaces. Similar set-ups could be used to locally evaluate AI systems for ‘social compatibility’, i.e., accessibility, acceptability, and ability to meet local needs. Such an approach would need to ensure a representative sample: There is an inherent risk of bias when recruiting mainly the ‘digitally interested’, potentially worsening inequity.

#### Theme 2: Shared understanding

A key part of *equity through influence* is *shared understanding*; to achieve more equitable influence over the implementation process, there needs to be a consistent understanding of core concepts, such as *‘what is health (in-)equity?’* and *‘what is the goal of the given intervention?’*. Again, this mirrors implementation science frameworks such as normalization process theory ^44^. There were differing thoughts on the role of AI in primary care among the participants; whilst AI developers seem to consider their products as largely ‘agnostic’, socially passive tools (as reflected in the focus on increased efficiency/productivity), they are being implemented as part of a complex system with multiple sometimes conflicting goals, as emphasized by the academic interviewees among others. An interviewee at an AI company echoed this view of their product as socially inert, and that affecting inequity was beyond their reach:

> We have worked on how on how to make the system easier for patients with poor vision, for example, but we have not focused on the broader structural aspects, such as equality in-between women and men in usage. I think this is something that we all as doctors have in the back of our heads, however.
>
> #20, AI company 1

A parallel can be drawn to Han et al’s ^45^ ethnography on a critical care IT system which was found to be increasing mortality due to a discrepancy between the perceived usage conditions (by the developers and implementers) and the actual situation (namely a reliance on informal communication and routines that were not replicated in the IT system), or Greenhalgh et al’s ^38^ rapid ethnography on the failed implementation of home-based health-IT for geriatric patients. In both cases, implantation failure arose from a discrepancy in how the producing and implementing stakeholders assumed that the products would be used and for which goals, and the reality of the users’ context. In some cases, the concept of health equity (equality in health outcomes) is in conflict with outright utilitarianism (the greatest good for the most patients), which can create tensions in designing and implementing interventions:

> A lot of health is built off a utilitarian view: the greatest good for the greatest number. If you’re following that, if you’re truly following that, then you’d say, well, actually, if it caused minor harm to a few people, if it caused greater good for a lot of people, then you should probably go with it.
>
> #9, GP and AI researcher

Clearly then, there is a link to the previously discussed theme *equity through influence*; in order to design and implement and AI system concordant with the NHS goal of reducing inequity in and through primary care, all stakeholders need to speak the same language, sharing experiences and creating a shared understanding of the context, the goals and the intervention.

#### Theme 3: Shared frameworks and collaborative platforms

In order to develop such a *shared understanding*, there needs to be standards and platforms in place: *shared frameworks.* This refers to both ways and regulations around sharing data as well as organizational structures to bring in local perspectives and feedback into commissioning, production, and regulation. Algorithmic impact assessments provide tools for a standardized and relatively convenient assessment of AI-products by commissioning and evaluating stakeholders. Current examples are rare but exist, e.g., one is being trialed by Ada Lovelace Institute at the moment ^46^. For equity improvement, future developments need a stronger and more detailed focus on equity and could draw from equity assessment tools such as the Health Equity Assessment Toolkit (HEAT ^47^):

> It’s almost irrelevant whether it’s a public sector or private sector or anything else: Using the HEAT is very much about understanding your audience, understanding the outcomes you want to achieve, and utilizing these principles by working with different professionals to get to the core of our audience and make a difference. That’s how I view it.
>
> #4, DHSC

Such AIAs could both act as a foundation for a common understanding and a practical guide for assessing AI products, before and after implementation. To develop such a tool, and to enable a *shared understanding* of how to address inequity, collaborative structures are needed. The AI and Digital Regulations Service (ADRS) is an example of a high-level collaborative system, while AHSNs provide potential starting points for more local entities. An improved data sharing framework is needed to enable said assessments and facilitate more equitable AI in the future (through access to real-life data on local populations), created in collaboration between regulatory organizations but presumably led by the ICO whilst maintaining privacy and subsequently trust:

> Data access is a big issue. The Goldacre review [A 2022 UK government-commissioned health-data whitepaper] advises to go close to the data source and do the research and development close to the implementation e.g., build systems at GP level. That enables easy trialing through running a system in parallel to real GP.
>
> #10, MHRA

#### Theme 4: Capacity building and knowledge generation

To accomplish the above and enable equitable implementation, *capacity building and knowledge generation* is needed across the whole ecosystem. This means different things for different stakeholders but highlights the need for resourcing and a coherent push for standards and understanding (e.g. how to assess an AI system from an equity perspective) and how to best conduct PPI for equitable AI, including increased capacity for assessment of new technology (e.g. MHRA), national evaluation (e.g. NICE), local assessment and evaluation (e.g. ICSs, primary care centers and networks) and through academia. An interviewed GP commented on the outlook for involving her practice in AI implementation:

> Am I super enthusiastic for that at the moment? No, because we don’t have the capacity. If as a cluster, we had funding for it and there was proper time to resource it, then yes, because you know, there’s a real chance that it might make a meaningful difference to patients and practices.
>
> #11, GP

In the primary care context, local ‘champions’ – highly motivated individuals or local organisations driving change on their own initiative – was mentioned as a way of achieving a collaborative and equitable implementation. However, such developments may also worsen inequity:

> There are the big centers where they’ve got a lot of money and a lot of expertise and there are places that people want to work. And basically, those are the places that are probably developing their technologies and adopting AI and being sort of mooted as the AI leaders across healthcare and the smaller trusts that struggle to recruit, that probably serve more deprived populations, have more challenges about providing healthcare and access to healthcare, and don’t have the money all the time or the expertise in order to develop these technologies or adopt and deploy them. I think they will just fall further and further behind.
>
> #14, clinician and AI researcher

This again emphasises the need for a high-level framework for AI implementation to promote equitable opportunities across localities with different circumstances.

#### Theme 5: Division of responsibility

Finally, the *division of responsibility* is discussed, depicted on the right side of the theory logic model (Figure 2). In the model, the stakeholders are presented as an organizational hierarchy of a typical AI intervention, from production to use. Different approaches to equity promotion are needed at different levels of the ecosystem, but (as depicted in the figure with a large double-sided arrow connecting thestakeholders with *equity through influence*), all stakeholders need to feed into a common understanding and approach. The one-sided arrow in the model depicts how the (inter-) national policy and regulation environment have a particular role to play in enabling *equity through influence, shared understanding,* and *shared frameworks*, as well as *capacity building and knowledge generation*.

In other words, high-level political and regulatory oversight is needed to accomplish the other themes, not by top-down action but though providing a clear set of regulations and protocols to enable a synchronized approach to be conducted at intermediate and local levels, in close collaboration between users, commissioners and producers. As described by an interviewee at MHRA:

> Ultimately it is very difficult for regulations to ensure equity. Local expertise is needed to assess it in the context [a product] is implemented. On the same note, MHRA has no agency to perform post-market surveillance; it is up the user to follow up.
>
> #10, MHRA

Meanwhile, local stakeholders such as ICSs, primary care centers (and -networks) and AHSNs needs to lead local implementation and involvement to produce interventions suited to the local needs, and subsequent evaluation; *applying* and *utilizing* the frameworks built by the higher system levels. Several interviewees talked about ‘place based’ implementation (as per NHS England an area of about 30 000 inhabitants ^48^), typically consisting of a large borough or a primary care network:

> I think practices are too small and the majority of ICSs I think are too big. What I want to do in North [City] is very different because that population has very little in common with East [City] population, for example. There are very few ICSs where they’ve got common issues across the whole economy; they tend to be in community-based sort of pockets. So, I think it has to be at a ‘place’ level.
>
> #6, ICS digital strategist

As mentioned, this is in line with the general ambition for ICSs to act as an umbrella and supporting organization for localized initiatives.

## Discussion

This paper brings together a set of organizational and public stakeholders in an ethnographically driven, explorative study. This approach, combined with the iterative constant-comparative methodology, allowed us to derive a theory from a broad range of empirical observations and create a uniquely holistic foundation for improvement of the AI implementation process for primary care, as set out in our key messages below. The study has several limitations: An inquiry assessing the intersection of AI, inequity and primary care will inevitably remain superficial to a degree, and this holistic system perspective is both a strength and weakness. Further, the study lacked demographic sampling for diverse participants among GPs and PwD groups. However, a ‘minimum’ demographic diversity criterion was applied to avoid an overly homogenous sample of PwD groups.

Through analysing the primary data against the frameworks outlined in the method section, a few key messages can be identified:

- The organizational ecosystem is largely unprepared for equitable implementation of AI in primary care. However, as mentioned the situation is multifaceted; strong systems for PPI, AIAs and a general movement towards localization of commissioning through ICSs and AHSNs is promising, but what is lacking is a unifying strategy encompassing the themes discussed above.
- High-level (inter-)national policy and regulation must promote frameworks for equitable implementation that enables all stakeholders, from developers to GPs and patients, to conduct implementation in an equitable manner. Such frameworks enable AIAs and implementation processes to be conducted at a local level, in turn enabling contextualization and local stakeholder involvement.
- Local capacity to conduct such processes subsequently need strengthening, and further research is needed to define how such AIAs and frameworks are best designed, including into means to effectively quantify and evaluate inequity effects of AI, and how to best involve the public in shaping AI interventions for health equity.

Whilst the difficulty of implementing technological interventions in complex systems is previously well studied ^13^ ^41^, the context of AI, inequity and primary care brings further complications. More than anything, the study highlighted the complexity of implementing AI in primary care, and the difficulties of assessing such processes from a inequity perspective. Part of the complexity stems from the relative absence of delineation; the term ‘AI in primary care’ as used here is very wide (as outlined in the introduction section). More pragmatically aimed future research will naturally have to narrow the definition to certain AI interventions or well-defined groups, to clearly be able to lay out the *mechanism of change* of a given intervention and the potential equity effects.

AI interventions in patient-facing healthcare will remain highly complex and will need to be studied in a manner that takes the sociotechnical complexity into account. Sociotechnical approaches such as ethnographically inspired inquiry certainly have an important role to play in future explorations.

## Data Availability

All data produced in the present study are available upon reasonable request to the authors, with the exception of identifiable interview and participant data.

## Acknowledgement

We acknowledge the valuable contributions by the two public advisors, Irum Durrani and Adele Thomas, as outlined above.

## Funding

This study was conducted as part of the PhD project “Artificial Intelligence and Health Inequities in Primary Care”, by Alexander d’Elia. The PhD project is funded by Applied Research Collaboration Northwest Coast (ARC NWC), in turn funded by the UK National Institute for Health Research (NIHR).

## Declaration of interests

The funder or the authors have no conflicts of interest to declare.

## Appendix 1

Conceptual framework model for how AI could affect inequities in primary care, forming part of the theoretical foundation for this study. From d’Elia et al (2022).

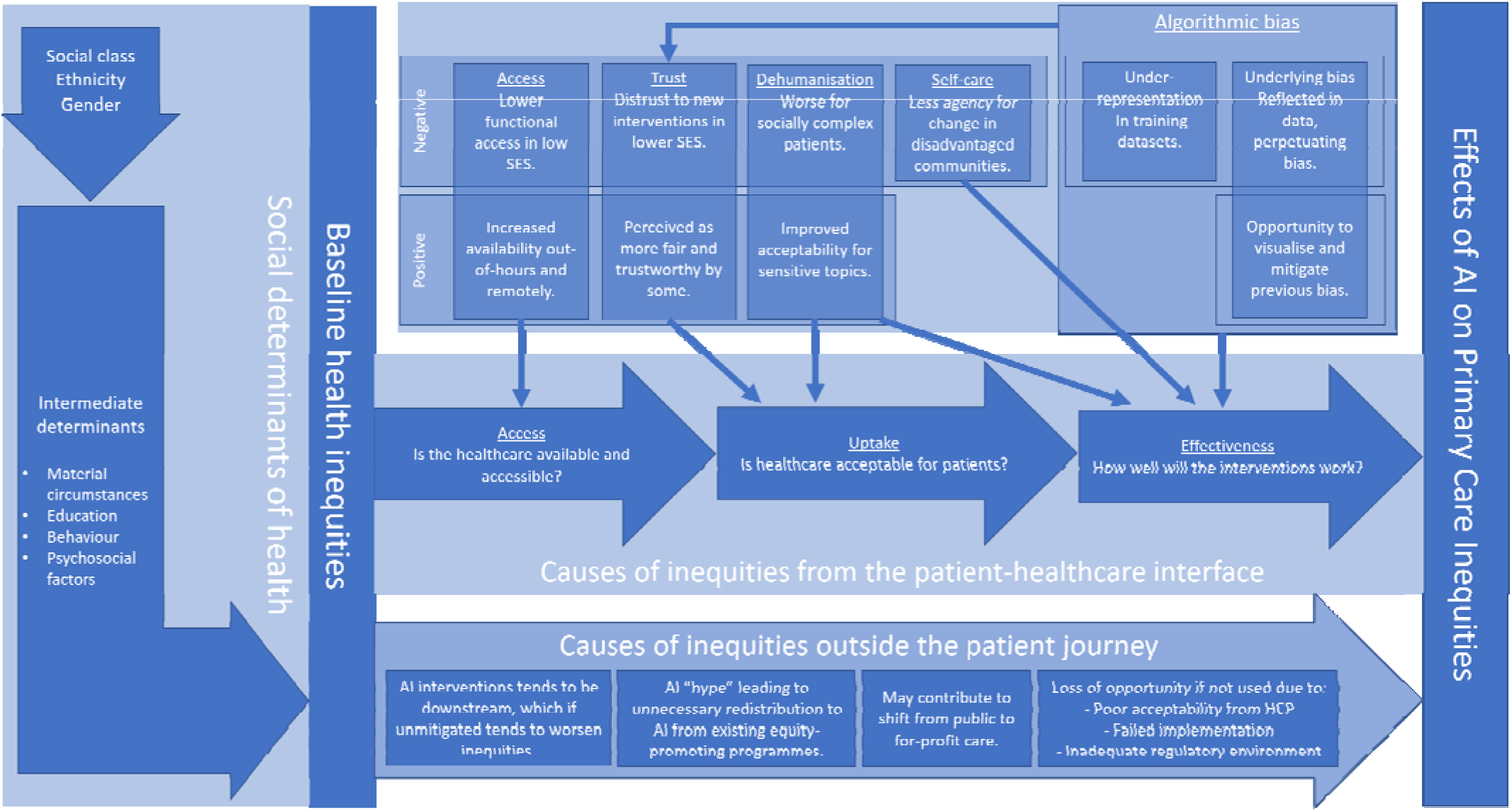

## Appendix 2

Schematic implementation process for AI in English NHS primary care, based on cited documents and fact-checked with interviewees:

1. Development:

a. A producer e.g. a private company, NHS organisation, academia, or any combination thereof (but most likely a private company) develops a product they believe is suitable for primary care.
b. The producer may or may not consult guidance on how to create an equity-improving AI product, for example through the abovementioned HEAT, through an AIA such as the one by The Ada Lovelace Institute ^46^, NICE ESF ^49^ or the guidance issued by The UK Office for Artificial Intelligence and The Alan Turing Institute ^20^.
2. Approval:

a. Producer seeks MHRA approval. MHRA produces standards according to UK MDR 2002 legislation. These standards are applied by accredited third-party organisations, which provides MHRA approval for the product. If it is a class 1 product (only advising) the producer can self-assess and provide documentation, otherwise the MHRA must be involved in the clinical assessment of efficacy.
b. If the system is intended to be used for traditionally primary-care-supplied services but not be used in actual NHS primary care (e.g. private healthcare or online symptom checkers), no further assessment is necessary.
c. If produced together with a local partner such as a university or a PCN, an AHSN or ICB may be involved in this process.
d. The producer may apply for the product to be place in the GPIT Futures catalogue of endorsed software. This is a not an absolute requirement for being procured into the NHS primary care system, but advised ^50^.
3. Procuring:

a. An ICB or a PCN (or rarely, a single primary care practice) sees a use case for the product and investigates procuring it. An AHSN is likely involved with the ICB in the case of ICS-wide employment. The ICS and the AHSN may have a PPI component advising the ICB on if and how to implement the product. How exactly the PPI is shaped varies between organisations, but it is expected (if not statutory) for both ICBs and AHSNs.
b. If the buyer is a PCN they would be expected, but not required, to ask the ICS and associated AHSN for support.
c. If under £40,000 and under twelve months contract time the buyer can self-procure, otherwise it has to be a competitive process between the various products meeting the specification in the GPIT Futures catalogue ^51^.
4. Operationalisation:

a. NHS Digital publishes guidance on operationalisation of new products. Their guidance is strictly operational in character and does not cover PPI/needs assessment and feedback from patients or end-users ^51^. However, it is assumed that those aspects have been covered to an extent by the ICB or associated AHSN.
5. Evaluation:

a. There are no detailed guidelines for evaluation, and no equity-specific evaluation is regularly done, but the buyer would in practice evaluate the products against the objectives it was brought in under. NHS Digital recommends annual evaluations ^50^. The CQC has a responsibility to assess whether products are *used* in a safe and effective manner from a clinical perspective but does not evaluate the effectiveness and other properties of the products themselves (e.g. algorithmic biases).

## Appendix 3

Based on the mapping in appendix 2 we combined fundamentals of implementation science (e.g. the need for a shared understanding of goals, benefits and measurables of a given intervention ^44^) and general approaches to inequity improvement in healthcare (such as HEAT ^47^, inequityAT ^52^ or Innov8 ^53^, among others). Our resultinga conceptual model for equitable implementation was developed to show how equitable implementation is possible. Here we illustrate the implementation chain that we recommend is followed from production to utilisation:

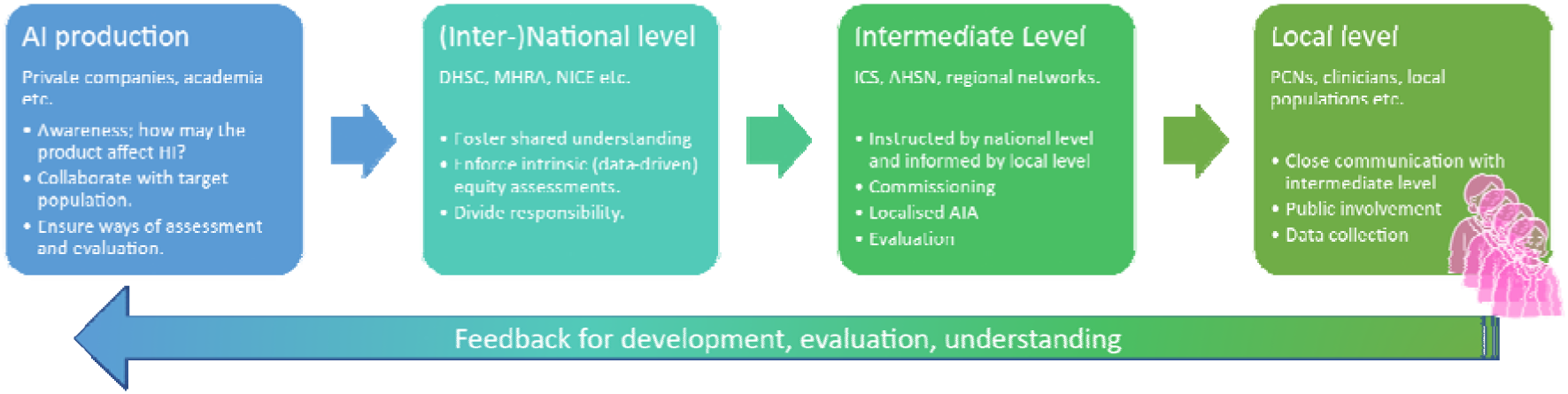

## Appendix 4

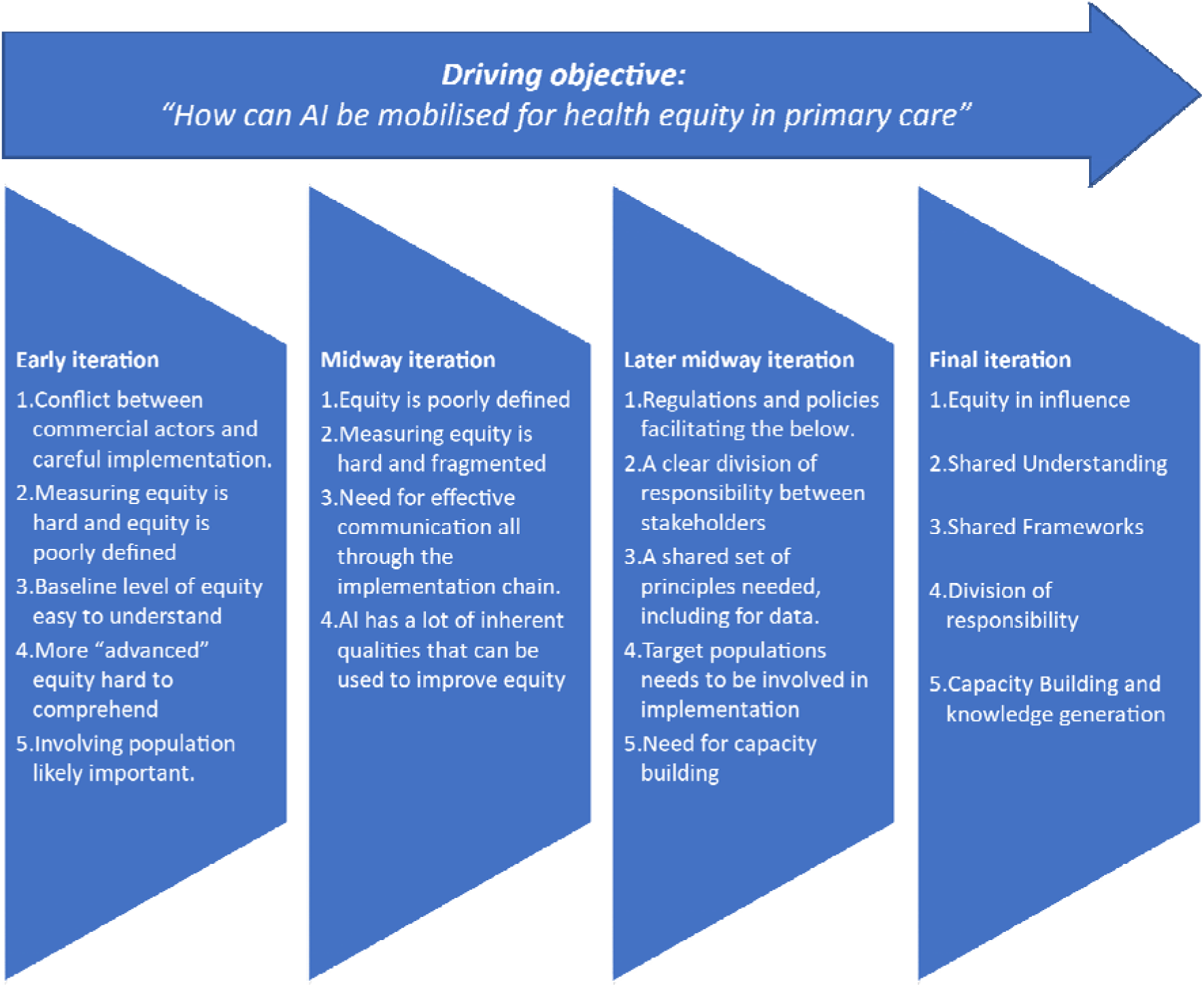

Sample of iterations of themes derived from coding interview data. Note that multiple minor revisions were made between these versions.

## Notes

### Competing Interest Statement

The authors have declared no competing interest.

### Author Declarations

The research ethics committee at The University of Liverpool gave ethical approval for this work, reference number 10229.

